# Accuracy of ICD-10 codes for suicidal ideation and action in pediatric emergency department encounters

**DOI:** 10.1101/2024.07.23.24310777

**Authors:** Rena Xu, Louisa Bode, Alon Geva, Kenneth D. Mandl, Andrew J. McMurry

## Abstract

**Objectives:** According to the ideation-to-action framework of suicidality, suicidal ideation and suicidal action arise via distinct trajectories. Studying suicidality under this framework requires accurate identification of both ideation and action. We sought to assess the accuracy of ICD-10 codes for suicidal ideation and action in emergency department (ED) encounters.

**Methods:** Accuracy of ICD-10 coding for suicidality was assessed through chart review of clinical notes for 205 ED encounters among patients 6-18 years old at a large academic pediatric hospital between June 1, 2016, and June 1, 2022. Physician notes were reviewed for documentation of past or present suicidal ideation, suicidal action, or both. The study cohort consisted of 103 randomly selected “cases,” or encounters assigned at least one ICD-10 code for suicidality, and 102 propensity-matched “non-cases” lacking ICD-10 codes. Accuracy of ICD-10 codes was assessed using sensitivity, specificity, positive predictive value (PPV), and negative predictive value (NPV).

**Results:** Against a gold standard chart review, the PPV for ICD-10 suicidality codes was 86.9%, and the NPV was 76.2%. Nearly half of encounters involving suicidality were not captured by ICD-10 coding (sensitivity=53.4%). Sensitivity was higher for ideation-present (82.4%) than for action-present (33.7%) or action-past (20.4%).

**Conclusions:** Many cases of suicidality may be missed by relying on only ICD-10 codes. Accuracy of ICD-10 codes is high for suicidal ideation but low for action. To scale the ideation-to-action model for use in large populations, better data sources are needed to identify cases of suicidal action.

## INTRODUCTION

Suicidality among youth, encompassing suicidal ideation, intentional self-harm, and suicide attempt,^1^ increased at an alarming rate during the COVID19 pandemic^2^ and remains an urgent public health concern. Accurate identification of suicidality is essential for research, allocation of healthcare resources, and development of public health programs and clinical interventions. Using International Classification of Diseases, 10th Revision (ICD-10) codes, which are designed primarily to support billing and reimbursement, can underestimate suicidality.^3–5^ A systematic review of 34 studies reported 2-19% sensitivity and 83-100% positive predictive value (PPV) for ICD-10 codes for suicide attempt and 43-88% PPV for ICD-10 codes for suicidal ideation among adolescents.^1^

Prevailing theories of suicide, including the three-step theory (3ST),^6^ interpersonal theory (IPTS), and integrated motivational-volitional model (IMV), are underpinned by an “ideation-to-action”^7^ framework. Under this framework, suicidal ideation and suicidal action are associated with distinct risk factors, and progression from ideation to action, which occurs in only a minority of individuals with ideation, depends on capability for suicide.^7–10^ The ideation-to-action model has been used in studies to identify risk factors for suicidal ideation versus action, as well as in interventions aimed at suicide prevention,^11^ but further research is needed to compare populations experiencing only ideation with those progressing to suicidal action to understand trajectories of progression. Expanded adoption of the ideation-to-action model hinges on accurate, consistent identification of each suicidality subtype. Health care claims are a frequently used data source for suicidality research, but prior studies of ICD-10 coding accuracy have not distinguished ideation and action sufficiently to allow study of the ideation-to-action framework. Understanding ICD-10 coding accuracy for suicidal ideation versus action is important to determine whether suicidality progression can be reliably identified in electronic health record (EHR) and payor claims data and whether such data can be used for care, research, and surveillance under the ideation-to-action model.

We sought to assess the accuracy of ICD-10 codes for capturing both suicidal ideation and action in emergency department (ED) encounters.

## MATERIALS AND METHODS

This retrospective study included all pediatric patients 6-18 years old presenting to the emergency department (ED) of a large, academic, tertiary care children’s hospital between June 1, 2016, and June 1, 2022. If patients required admission to the hospital from the ED, the admission was considered part of the same encounter.

Encounter data were extracted from the EHR, including patient demographics, diagnoses, and clinical notes. Patient demographics included sex, age at visit, and patient-reported race. The ED provider notes and, if present, psychiatry consultation notes and inpatient discharge summaries were also included.

ICD-10 codes for suicidality were identified from prior studies.^2,12–20^ “Cases” were defined as encounters including one or more of these suicidality ICD-10 codes (**Supplemental file 1**). “Non-cases” were defined as encounters in the same period without ICD-10 codes for suicidality. Non-cases were selected using propensity score matching (PSM), a technique commonly used to control for confounding. The PSM model was fit using patient demographics, ICD-10 diagnoses, and number of documented encounters as independent variables and presence of a suicidality ICD-10 code as the dependent variable (**Supplemental file 2**). The PSM model outputs both the effect sizes for each variable and the likelihood that the patient encounter has a diagnosis of suicidality (propensity score).

Consistent with the established “ideation-to-action” model, suicidality subtypes were defined in this study as suicidal ideation or suicidal action, with the latter including both intentional self-harm (non-suicidal self-injury) and suicide attempt. Suicidality progression was dichotomized as past or present to distinguish current suicidality from a history of suicidality disclosed in the present encounter. This resulted in four suicidality subtypes: *ideation-past, ideation-present, action-past, action-present*. Each encounter could be assigned between zero and four suicidality subtypes.

Guidelines for chart review were developed to classify each subtype of suicidality (**Figure 1**, and additional details in **Supplemental file 3**). Two reviewers (RX, AM) collectively reviewed a total of 205 encounters, consisting of 103 cases and 102 propensity-matched non-cases. Reviewers were blinded as to whether encounters were cases or non-cases. Following an initial independent review of 50 encounters, inter-reviewer agreement was assessed using Cohen’s kappa (κ)^21–23^. Instances of disagreement were adjudicated by a third member of the study team (AG) and discussed by reviewers to resolve any disagreements. Reviewers then reviewed 155 additional unique encounters (64 by RX, 91 by AM).

**Figure 1.**
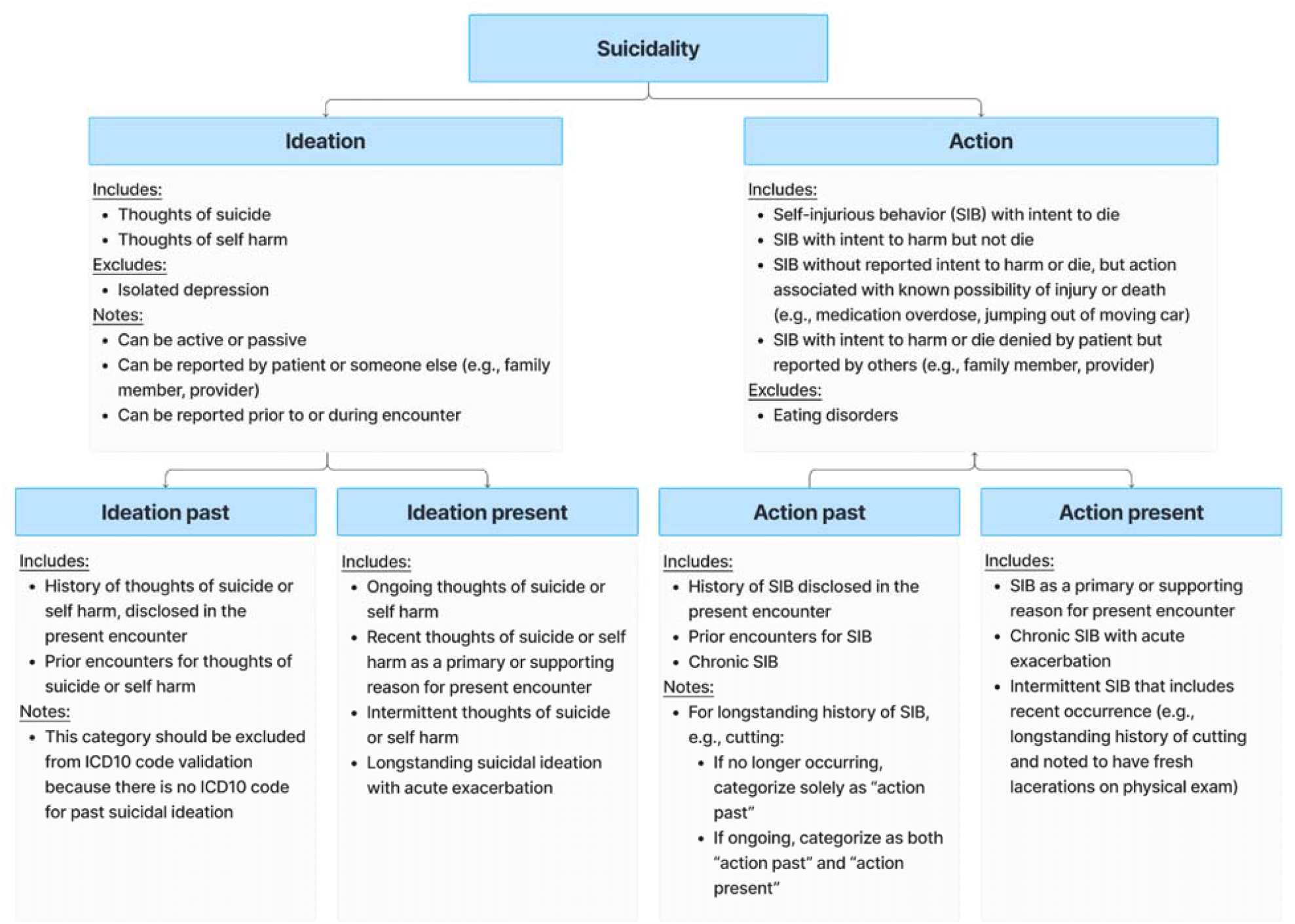
Summary of chart review guidelines for suicidality subtypes.

Agreement between classification by reviewers and by ICD-10 coded diagnoses was assessed across all 205 reviewed encounters. Sensitivity, specificity, PPV, and NPV were calculated for *ideation-present, action-past, and action-present (but not ideation-past* since ICD-10 does not include a corresponding code).

## RESULTS

The overall ED population comprised 59,866 patients aged 6-18 years old with 90,980 ED encounters during the study period (**Table 1**). Just over half of ED encounters were for children aged 6-11 years old (51.9%). Slightly over half of encounters (51.7%) were for male patients. ICD-10 codes for suicidality were present in 2,637 ED encounters among 2,230 patients, for an ICD-10-based suicidality prevalence of 2.9%. Most suicidality encounters were for adolescents aged 12-18 years (88.2%) and females (69.2%). The distribution of assigned ICD-10 codes among suicidality encounters was suicidal ideation 86.2%, self-harm 39.1%, and suicide attempt 6.7%.

**Table 1.** Characteristics of overall ED population and study cohort.

|  | **A) Overall population** |  | **B) Study cohort** |  |
| --- | --- | --- | --- | --- |
| **Demographic** | **All ED encounters**<br>**(% of total)** | **ED encounters**<br>**with ICD-10 code**<br>**for suicidality**<br>**(% of total)** | **Cases**<br>**(% of total)** | **Non-cases**<br>**(% of total)** |
| **Total** | **90,980 (100%)** | **2,637 (100%)** | **103 (100%)** | **102 (100%)** |
| Age 6-11 yrs | 47,253 (51.9%) | 310 (11.8%) | 17 (16.5%) | 14 (13.7%) |
| Age 12-18 yrs | 43,727 (48.1%) | 2327 (88.2%) | 86 (83.5%) | 88 (86.3%) |
| Male | 47,045 (51.7%) | 812 (30.8%) | 25 (24.3%) | 39 (38.2%) |
| Female | 43,929 (48.3%) | 1825 (69.2%) | 78 (75.7%) | 63 (61.8%) |

A) Overall population refers to all ED encounters in the hospital during the study period (June 1, 2016, to June 1, 2022). B) Study cohort refers to cases and non-cases that were reviewed to assess accuracy of ICD-10 coding for suicidality.

As compared to overall ED encounters with ICD-10 code for suicidality, cases randomly sampled for chart review (N=103) as well as non-cases identified via propensity-score matching (N=102) had similar demographic proportions (p>0.05 for both age and gender on chi-square analysis; **Supplemental file 4**).

Overall agreement between chart reviewers was almost perfect (κ=0.862), with agreement by suicidality subtype ranging from almost perfect for ideation-present (κ=0.959) and action-past (κ=0.830) to substantial for action-present (κ=0.747). The PPV of ICD-10 suicidality codes was high (PPV=86.9%) (**Table 2**). However, ICD-10 codes failed to capture roughly half of suicidality cases identified by reviewers (sensitivity=53.4%). Among propensity-matched non-cases, nearly a quarter of encounters represented false negatives (NPV=76.2%) — i.e., chart review indicated suicidality, but ICD-10 codes were lacking.

**Table 2.** Accuracy of ICD-10 codes for suicidality by subtype.

| **Suicidality subtype** | **Sensitivity** | **Specificity** | **PPV** | **NPV** |
| --- | --- | --- | --- | --- |
| Overall | 0.534 | 0.953 | 0.869 | 0.762 |
| Action-past | 0.204 | 0.954 | 0.658 | 0.708 |
| Action-present | 0.337 | 0.985 | 0.878 | 0.802 |
| Ideation-present | 0.824 | 0.891 | 0.91 | 0.791 |

“Overall” agreement for suicidality includes *action-past*, *action-present*, and *ideation-present*.
By subtype, ICD-10 accuracy was highest for *ideation-present* and lowest for *action-past*.

Differences in ICD-10 code accuracy were observed by suicidality subtype. The PPV of ICD-10 codes was higher for *ideation-present* (91%) as compared to either *action-present* (87.8%) or *action-past* (65.8%). ICD-10 codes captured most encounters with present suicidal ideation (sensitivity=82.4%) but only a third of all encounters documenting present action (sensitivity=33.7%) and even fewer encounters documenting past action (sensitivity=20.4%).

Detailed chart review guidelines and results are included in **Supplemental files 3 and 4**, respectively.

## DISCUSSION

Growing rates of suicidality ^21–24^ among youth in the U.S. heighten the urgency to understand drivers of suicidal ideation and progression from ideation to attempt. Consistent with prior studies,^1^ ICD-10 coding are reliable for identifying suicidal ideation among youth.

However, we find they do not accurately identify action. While the predictive value of ICD-10 codes for suicidality was high in this study, nearly half of all encounters that involved suicidality lacked corresponding ICD-10 codes.

Most studies to date have focused on PPV^1,25,26^ and have not assessed the rate of false negatives --i.e., encounters that involved suicidality but were not assigned an ICD code for suicidality. Because propensity score matching was used to identify non-cases with the greatest probability of involving suicidality, targeted review of charts could be performed to approximate NPV and gain a more comprehensive view of coding accuracy. The results suggest the true prevalence^27^ of suicidality in the evaluation cohort was higher than what was calculated using ICD-10 codes.

Accurate data are needed to use the ideation-to-action conceptual model in care, research, and surveillance; to allocate resources appropriately; and to implement interventions. To our knowledge, this study is the first to adopt an ideation-to-action framework for chart review and assessment of coding accuracy. Limitations of the study include its retrospective design, restriction to a single site, and sole focus on ED visits. Furthermore, this study did not assess possible varying accuracy of ICD-10 codes across sociodemographic groups. Additionally, the PSM model did not account for frequency of inpatient admission from the ED or total length of stay. It is possible that patients who spent more time in the hospital, whether due to an admission or prolonged observation in the ED, were exposed to more opportunities for accurate ICD-10 coding. The study may also underestimate the real-world NPV of ICD-10 codes, since the PSM model selected for non-cases that were most likely to include suicidality missed by ICD-10 codes.

Future research may further elucidate ICD-10 coding accuracy for suicidality. For example, accuracy may vary over time for individuals with multiple ED encounters for suicidality. Bentley et al.^28^ recently used single-system EHR data to assess the accuracy of ICD codes for patients with multiple suicide attempts and reported a PPV of 90% for distinct encounter codes entered at least five days apart. There may be a difference in ICD-10 code sensitivity for capturing suicidal action in patients who have had multiple prior encounters for suicidality as compared to individuals presenting with suicidal action for the first time. Future studies could help to assess the reliability of ICD-10 coding for mapping progression from suicidal ideation to action and more broadly from subacute mental illness to acute suicidality. Future work should also include automated methods for processing clinical notes. Advances in natural language processing will allow for extraction of computable phenotypes of suicidality. This could facilitate replication of the present study across multiple sites of care^29–32^ and provide a more accurate source of data on suicidality as compared to ICD-10 codes.

## Data Availability

All data produced in the present work are contained in the manuscript

## ACKNOWLEDGMENTS

We thank our colleagues at the Massachusetts Department of Public Health for providing public health insights into the challenges of suicidality progression in children. The authors especially thank Sita Smith, Rosa Ergas, Alyssa Anwar, Matthew Tumpney, Dylan Keating, Lauren Larochelle, Victoria Nielsen, Lindsey Sagasta, Megha Parikh, Jennifer Ledet, and Anna Hammelrath, as well as valued colleagues at the CDC foundation, Elizabeth Sprouse and Lyndsey Kirchner. We also thank Tim Miller and Karen Olson for their advice on the study methodology.

## INSTITUTIONAL REVIEW BOARD STATEMENT

The Institutional Review Board of Boston Children’s Hospital waived ethical approval for this work (IRB-P00043211).

