## Supplemental File 2 for "Accuracy of ICD-10 codes for suicidal ideation and action in pediatric emergency department encounters"

**Supplemental File 2: Propensity Score Matching**

**Contents:**

**Table S2:** **Propensity score matching variables**

**Figure S2: Effect size of variables before and after propensity matching**

Cases were matched to non-cases using PsmPy (<https://pubmed.ncbi.nlm.nih.gov/36086543/>) an open-source Python software for propensity score matching. PsmPy was configured to use logistic regression and k-nearest neighbors without a caliper width. The logistic regression output score denotes the propensity (probability) that each encounter matches the suicidality case definition. The PsmPy input variables included demographics (gender, age group, patient reported race); documented healthcare utilization; and DSM-5 diagnostic categories. Study period and selection criteria are described in the Methods section of the manuscript. History was trimmed such that future encounter diagnosis would not influence prior predicted scores. The PsmPy logistic regression classifier was used to score the probability that each encounter would include an ICD-10 diagnosis code for suicidality. Cases of suicidality were then 1:1 matched to non-cases based on the PsmPy predicted propensity scores using k-nearest neighbors.

**Table S2. Propensity score matching variables**

| **Type** | **Variable** | **Description** |
| --- | --- | --- |
| Demographics | Gender | Biological sex assigned at birth. |
| Demographics | Age Group | Patient age at visit. Age groups were children aged 6-11 years and adolescents aged 12-18 years. |
| Demographics | Patient reported race | Centers for Disease Control and Prevention (CDC) race codes   - American Indian or Alaska Native - Asian - Black or African American - Native Hawaiian or Pacific Islander - Other - White - Unknown |
| Utilization | Visit count, total | Number of patient encounters during the study period. |
| Utilization | Count discharge summaries | Number of documented hospital discharges. |
| Utilization | Count ED notes | Number of documented ED encounters. |
| Utilization | Count Psych Eval | Number of documented psychiatric evaluations. |
| Utilization | Count Psych Consult | Number of documented psychiatric consults. |
| Utilization | Person awaiting admission to adequate facility elsewhere | Binary variable: ICD-10 code Z75.1 |
| DSM-5 | Intellectual Disabilities | F70, F71, F72, F73, F88, F79 |
| DSM-5 | Communication Disorders | F80.9, F80.0, F80.81, F80.89, F80.9 |
| DSM-5 | Autism Spectrum Disorder | F84.0 |
| DSM-5 | Attention-Deficit/Hyperactivity Disorder | F90.0, F90.1, F90.2, F90.8, F90.9 |
| DSM-5 | Specific Learning Disorder | F81.0, F81.2, F81.81 |
| DSM-5 | Motor Disorders | F82, F98.4 |
| DSM-5 | Tic Disorders | F95.2, F95.1, F95.0, F95.8, F95.9 |
| DSM-5 | Other Neurodevelopmental Disorders | F88, F89 |
| DSM-5 | Schizophrenia Spectrum and Other Psychotic Disorders | F21, F22, F23, F20.81, F20.9, F25.0, F25.1, F06.0, F06.2, F28, F29, F06.1 |
| DSM-5 | Bipolar Disorders | F31.0, F31.1, F31.10, F31.11, F31.12, F31.13, F31.2, F31.3, F31.30, F31.31, F31.32, F31.4, F31.5, F31.6, F31.60, F31.61, F31.62, F31.63, F31.64, F31.7, F31.70, F31.71, F31.72, F31.73, F31.74, F31.75, F31.76, F31.77, F31.78, F31.8, F31.81, F34.0, F06.33, F06.34, F31.89, F31.9 |
| DSM-5 | Depressive Disorders | F34.8, F32.0, F32.1, F32.2, F32.3, F32.4, F32.5, F32.8, F32.81, F32.89, F32.9, F33.0, F33.1, F33.2, F33.3, F33.4, F33.40, F33.41, F33.42, F33.8, F33.9, F34.1, N94.3, F06.31, F06.32, F06.34, F32.8, F32.9 |
| DSM-5 | Anxiety Disorders | F93.0, F94.0, F40.218, F40.228, F40.230, F40.231, F40.232, F40.233, F40.248, F40.298, F40.10, F41.0, F40.00, F41.1, F06.4, F41.8, F41.9 |
| DSM-5 | Obsessive-Compulsive and Related Disorders | F42, F45.22, F63.2, L98.1, F06.8 |
| DSM-5 | Trauma- and Stressor-Related Disorders | F94.1, F94.2, F43.10, F43.0, F43.20, F43.21, F43.22, F43.23, F43.24, F43.25, F43.8, F43.9 |
| DSM-5 | Dissociative Disorders | F44.81, F44.0, F44.1, F48.1, F44.89, F44.9 |
| DSM-5 | Somatic Symptom and Related Disorders | F45.1, F45.21, F44.4, F44.5, F44.6, F44.7, F54, F68.10, F45.8, F45.9 |
| DSM-5 | Feeding and Eating Disorders | F98.3, F50.8, F98.21, F50.01, F50.02, F50.2, F50.9 |
| DSM-5 | Elimination Disorders | F98.0, F98.1, N39.498, R15.9, R32, R15.9 |
| DSM-5 | Sleep-Wake Disorders | G47.00, G47.10, G47.411, G47.419, G47.429, G47.33, G47.31, G47.37, R06.3, G47.37, G47.34, G47.35, G47.36, G47.20, G47.21, G47.22, G47.23, G47.24, G47.26, F51.3, F51.4, F51.5, G47.52, G25.81, G47.09, G47.00, G47.19, G47.10, G47.8, G47.9 |
| DSM-5 | Sexual Dysfunctions | F52.32, F52.21, F52.31, F52.22, F52.6, F52.0, F52.4, F52.8, F52.9 |
| DSM-5 | Gender Dysphoria | F64.1, F64.2, F64.8, F64.9 |
| DSM-5 | Disruptive, Impulse-Control, and Conduct Disorders | F91.3, F63.81, F91.1, F91.2, F91.9, F60.2, F63.1, F63.3, F91.8, F91.9 |
| DSM-5 | Substance-Related and Addictive Disorders | F10.10, F10.20, F10.129, F10.229, F10.929, F10.239, F10.232, F10.99, F15.929, F15.93, F15.99, F12.10, F12.20, F12.129, F12.229, F12.929, F12.122, F12.222, F12.922, F12.288, F12.99, F16.10, F16.20, F16.129, F16.229, F16.929, F16.983, F16.99, F18.10, F18.20, F18.129, F18.229, F18.929, F18.99, F11.10, F11.20, F11.129, F11.229, F11.929, F11.122, F11.222, F11.922, F11.23, F11.99, F13.10, F13.20, F13.129, F13.229, F13.929, F13.239, F13.232, F13.99, F15.10, F14.10, F15.20, F14.20, Z72.0, F17.200, F17.203, F17.209, F19.10, F19.20, F19.129, F19.229, F19.929, F19.239, F19.99, F63.0 |
| DSM-5 | Neurocognitive Disorders | F05, R41.0, F01.51, F01.50, G31.9, G31.84, F02.81, F02.80, R41.9 |
| DSM-5 | Personality Disorders | F60.0, F60.1, F21, F60.2, F60.3, F60.4, F60.81, F60.6, F60.7, F60.5, F07.0, F60.89, F60.9 |
| DSM-5 | Paraphilic Disorders | F65.3, F65.2, F65.81, F65.51, F65.52, F65.4, F65.0, F65.1, F65.89, F65.9 |
| DSM-5 | Other Mental Disorders | F06.8, F09, F99 |
| DSM-5 | Medication-Induced Movement Disorders and Other Adverse Effects of Medication | G21.11, G21.19, G21.0, G24.02, G25.71, G24.01, G24.09, G25.71, G25.1, G25.79, T43.205A, T43.205D, T43.205S, T50.905A, T50.905D, T50.905S |

Three variable types were included in the PSM model: demographics, healthcare utilization, and Diagnosis and Statistical Manual of Mental Disorders, Fifth Edition (DSM-5) category. There are 28 DSM-5 categories; each category is either true or false (dichotomous variables). ED = emergency department.

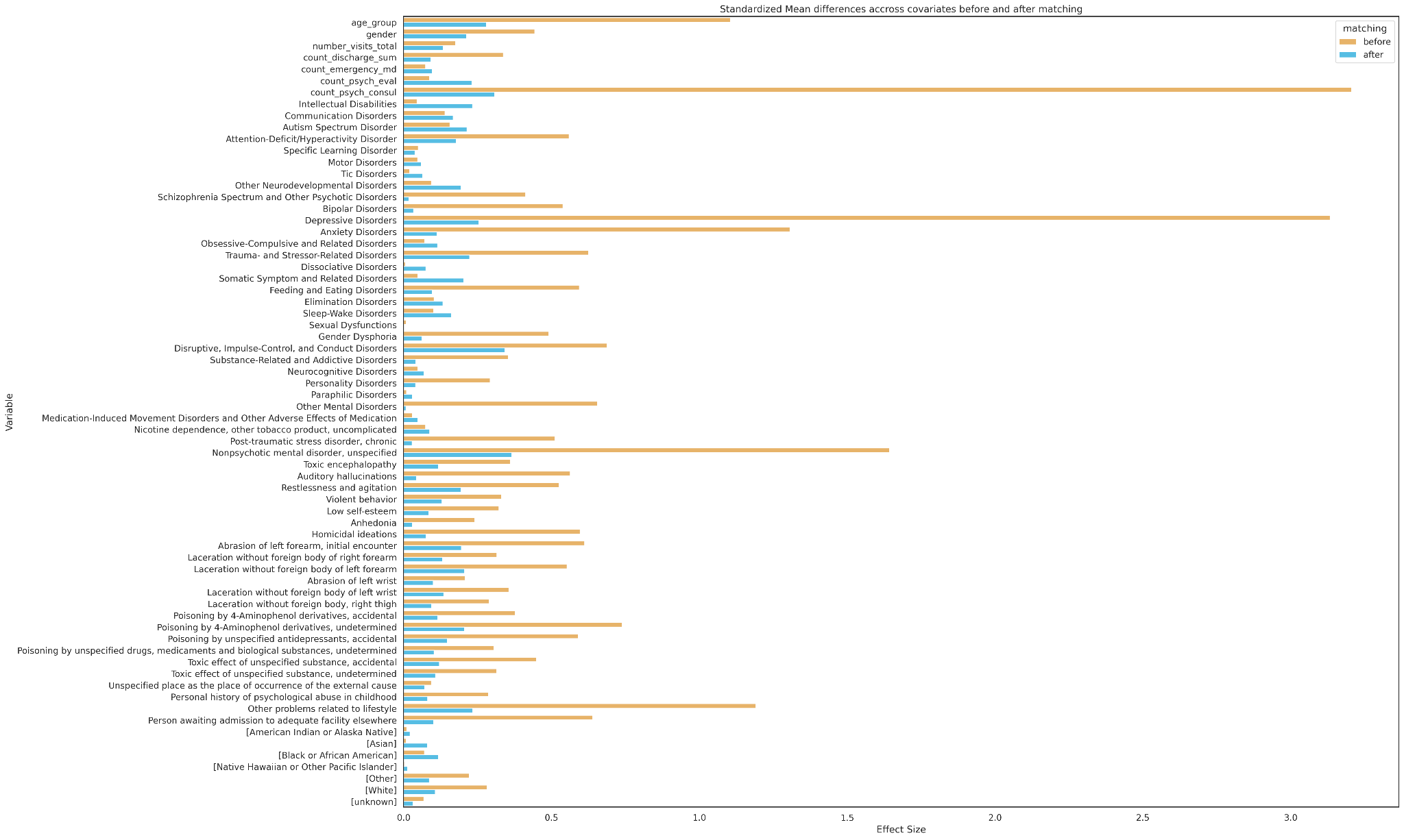

**Figure S2. Effect size of variables before and after propensity matching.** X-axis denotes effect size. Y-axis denotes each variable. Brown and blue bars denote variable effect size before and after matching, respectively. The largest effect size difference before matching was observed for the healthcare utilization variable “count psychiatric consults,” followed by the DSM-5 category “depressive disorders.”
