## Supplemental File 3 for "Accuracy of ICD-10 codes for suicidal ideation and action in pediatric emergency department encounters"

**Supplemental File 3: Suicidality Chart Review Guidelines**

**Contents:**

**Table S3.1:** **Note section criteria**

**Table S3.2: Suicidality subtype criteria**

**Table S3.3: Examples** **of suicidality subtype classification**

**Table S3.1. Note Section Criteria**

| **Note Section Heading** | **Include as past or present encounter** | **Exclude** |
| --- | --- | --- |
| Chief Complaint Reason for Visit Reason for Evaluation | Present |  |
| History of Present Illness | Past or present, as documented in section |  |
| Past Medical History | Past |  |
| Family History |  | Exclude suicidality of family members |
| Mental Status Exam,  PHQ-9, ASQ, other Psychiatric Questionnaires | Past or present, as documented in section |  |
| Assessment and Plan, Course, Evaluation | Past or present, as documented in section |  |
| Medical Decision Making | Past or present, as documented in section |  |
| Diagnosis |  | Exclude all codes for suicidality |
| Diagnostic Imaging Studies |  | Exclude (example: EKG, XR) |
| Laboratory studies |  | Exclude (example: UA, blood counts) |
| Other sections | Past or present, as documented in section |  |

Summary of note sections to include or exclude for chart review. **“**Note section heading” column lists examples of common section headings present within ED notes, psychiatry notes, and/or discharge summaries. “Include” column refers to whether the section should be included in chart review to categorize past and/or present suicidality. If the “include” column is blank, the section is always excluded during chart review. “Exclude” column clarifies why the section should be ignored and optionally provides examples. Across all note sections, ICD-10 codes and ICD-10 terminology are excluded from review if present.

**Table S3.2. Suicidality subtype criteria**

| **Subtype** | **Include** |
| --- | --- |
| **Ideation-past** | Ideation documented for previous healthcare visit or episode outside of healthcare setting. Typically “history of SI”, “chronic SI”, “worsening SI.”  Notice: ICD-10 does not contain any available codes for ideation-past. |
| **Ideation-present** | Ideation documented for current visit. Typically included in chief complaint, history of present illness, or physical exam as “mild to severe SI,” “worsening SI,” “thoughts of self-harm.” |
| **Action-past** | Self-harm or suicide attempt documented for previous encounter or episode outside of healthcare setting. Typically mentioned as “previous,” “second,” or “history of” self-harm or suicide attempt. |
| **Action-present** | Current encounter is for self-harm or suicide attempt, with or without history of suicidality. If the patient mentions self-harm or attempt, then action-present is true. If the provider assesses self-harm or attempt then it also true, even if the patient denies. |

Definitions for four suicidality subtypes: *ideation-past, ideation-present, action-past*, and/or *action-present*. More than one suicidality subtype may be documented in a single note.

**Table S3.3. Examples of suicidality subtype classification**

**Ideation-past**

| **Examples** | **Rationale** |
| --- | --- |
| - History of SI - Disclosed suicidal thoughts to school guidance counselor last semester - Disclosed thoughts of hurting himself to school guidance counselor last semester - Told parents he wished he was dead (in setting of previous encounter) - Thoughts started after traumatic life event | Ideation: thoughts of suicide or self-harm   Past: thoughts disclosed prior to the current encounter |

**Ideation-present**

| **Examples** | **Rationale** |
| --- | --- |
| - At risk of self-injurious behavior - Consult Request: concern for self-harm.  Pt denies suicidality. | Ideation: thoughts of suicide or self-harm   Present: “at risk”, “concern for” |
| - Chief Complaint: suicidal ideation - HPI: Presents with SI - Recent SI related to present reason for visit - Pt here with me today with SI - Ongoing SI with plans and intent - Suicide pact with friends - Phone text messages plan to hang herself | Ideation: SI documented   Present: “presents with” |
| - Here today with worsening SI | Ideation: encounter is for SI  Present: “worsening SI” implies acute ideation |
| - Patient thinks about what it would be like to be dead. - Intermittent thoughts about suicide - Thoughts about hurting myself - Passive SI with no plan | Ideation: thinks about self-harm and/or death  Present: “passive,” ”intermittent” |
| - Suicidal Ideation: 1 = Very Mild - Suicidal Ideation: 2, 3, 4, or 5 - Suicidal Ideation: 3 = Moderate - Suicidal Ideation: 6= Extremely severe | Ideation: Exam score  Present: Exam result at time of visit |

**Action-past**

| **Examples** | **Rationale** |
| --- | --- |
| - Pt tried to kill himself with random pills from the family medicine cabinet when his mother died. Pt was admitted to a psych facility for over a week before being discharged. Things at home settled for a while until recently when SI returned and he started cutting again last month. - Last year attempted to act on suicidal intent by holding a knife to his chest before family member stopped them | Action: suicide attempt.  Past: history of attempt before today (present encounter). |
| - Tried drowning 3 days ago (prior ED visit) - Recent suicide attempt one month ago - Previously attempted to asphyxiate with nitrous. - Multiple suicide attempts over the last year. - “History of” self-harm - “History of” cutting himself - “Chronic” self-harming behavior - PMH: previously diagnosed with self-harming behaviors - Cut in the past starting in December and most recently cut 1 month ago. Pt does not show any marks. - Pt reports self-cutting over a year ago for stress relief. Pt denies self-harm presently. No visible scars at time of observation. - History of cutting, last episode was 2020. | Action: self-harm (with or without intent to die)  Past: Previous time or ED visit |
| - HPI: presents today for SIB. Self-cutting for over a year, progressively getting worse. | Action: SIB  Present: presents today   Past: progressively getting worse  Note: code as both action-past and action-present. |

**Action-present**

| **Examples** | **Rationale** |
| --- | --- |
| - HPI: presents today for SIB. Self-cutting for over a year, progressively getting worse. | Action: SIB  Present: presents today   Past: progressively getting worse  Note: code as both action-past and action-present. |
| - CC: I cut myself so my therapist told me to go to the hospital - CC: self-harm with melatonin | Action: patient stated self-harm  Present: chief complaint section |
| - HPI: self-mutilation by knife - HPI: patient reports self-harm by razorblade.  Pt did not intend to die. - HPI: patient denies self-harm, contrary ED care provider opinion. - Suicidal/self-injurious behaviors: none reported by pt, but mother shared recent SIB | Action: self-harm, even if the patient denies.  Present: HPI (history of present illness). Stated in current tense. |
| - Forearms show superficial lacerations (in the context of SI/ideation) | Action: physical evidence of self-harm, even if the patient did not mention.   Present: cuts appear fresh on exam |

Examples of suicidality subtype classification. In the “examples” column, each bullet point represents an excerpt from reviewed clinical notes. The “rationale” column explains why each example was classified as action versus ideation and past versus present.
