## Supplemental File 4 for "Accuracy of ICD-10 codes for suicidal ideation and action in pediatric emergency department encounters"

**Supplemental File 4: Additional Statistical Measures**

**Contents:**

**Table S4.1: Chi square analysis for chart review population**

**Table S4.2: Agreement between reviewers by suicidality subtype**

**Table S4.3: Accuracy of ICD-10 codes by reviewer**

**Table S4.1. Chi square analysis for study population demographics**

|  | **A) Cases (N=103)** | | **B) Non-cases (N=102)** | |
| --- | --- | --- | --- | --- |
| **Demographic** | ***X*^2^** | **P-value** | ***X*^2^** | **P-value** |
| Age group 6-11 vs. 12-18 yrs | 2.13 | 0.14 | 0.37 | 0.55 |
| Male vs. Female | 1.99 | 0.16 | 2.54 | 0.11 |

Age and gender proportions did not differ significantly for A) chart-reviewed cases or B) chart-reviewed non-cases as compared to all ED encounters with ICD-10 code for suicidality. P<0.05 was considered significant.

**Table S4.2. Agreement between reviewers by suicidality subtype**

| **Suicidality subtype** | **Sensitivity** | **Specificity** | **PPV** | **NPV** |
| --- | --- | --- | --- | --- |
| Overall | 0.957 | 0.915 | 0.909 | 0.96 |
| Action-past | 0.895 | 0.935 | 0.895 | 0.935 |
| Action-present | 0.917 | 0.895 | 0.733 | 0.971 |
| Ideation-past | 1 | 0.812 | 0.919 | 1 |
| Ideation-present | 0.966 | 1 | 1 | 0.955 |

Agreement between two reviewers (AM, RX) for suicidality by subtype. PPV and NPV are positive and negative predictive values, respectively. “Overall” agreement between reviewers includes *action-past*, *action-present*, *ideation-past*, and *ideation-present*.

**Table S4.3. Accuracy of ICD-10 codes by reviewer**

A) **Reviewer 1 (RX)**

| **Suicidality subtype** | **Sensitivity** | **Specificity** | **PPV** | **NPV** |
| --- | --- | --- | --- | --- |
| Overall | 0.494 | 0.944 | 0.89 | 0.669 |
| Action-past | 0.14 | 0.953 | 0.7 | 0.587 |
| Action-present | 0.316 | 0.987 | 0.923 | 0.743 |
| Ideation-present | 0.816 | 0.842 | 0.912 | 0.696 |

B) **Reviewer 2 (AM)**

| **Suicidality subtype** | **Sensitivity** | **Specificity** | **PPV** | **NPV** |
| --- | --- | --- | --- | --- |
| Overall | 0.573 | 0.961 | 0.848 | 0.855 |
| Action-past | 0.267 | 0.955 | 0.615 | 0.828 |
| Action-present | 0.357 | 0.982 | 0.833 | 0.86 |
| Ideation-present | 0.831 | 0.939 | 0.907 | 0.885 |

Accuracy of ICD-10 codes for suicidality based on chart review by A) reviewer 1 and B) reviewer 2. “Overall” agreement includes *action-past*, *action-present*, and *ideation-present*.
